# Data-driven extraction of unstructured electronic health records to evaluate glioblastoma treatment patterns

**DOI:** 10.1101/2023.04.25.23289047

**Authors:** Akshay Swaminathan, Alexander L. Ren, Janet Y. Wu, Aarohi Bhargava-Shah, Ivan Lopez, Ujwal Srivastava, Vassilis Alexopoulos, Rebecca Pizzitola, Brandon Bui, Layth Alkhani, Susan Lee, Nathan Mohit, Noel Seo, Nicholas Macedo, Winson Cheng, William Wang, Edward Tran, Reena Thomas, Olivier Gevaert

**Affiliations:** Stanford University School of Medicine, Stanford, CA; Cerebral Inc., San Francisco, CA; Department of Computer Science, Stanford University, Stanford, CA; Department of Electrical Engineering; Stanford University, Stanford, CA; Department of Symbolic Systems, Stanford University, Stanford, CA; Department of Human Biology, Stanford University, Stanford, CA; Department of Materials Science and Engineering, Stanford University, Stanford, CA; Department of Psychology, Stanford University, Stanford, CA; Department of Computer Science, Human Biology, Stanford University, Stanford, CA; Department of Sociology, Stanford University, Stanford, CA; Department of Biology, Stanford University, Stanford, CA; Department of Radiology, Stanford University School of Medicine, Stanford, CA; Department of Chemistry, Department of Computer Science, Stanford University, Stanford, CA; Department of Bioengineering, Stanford University, Stanford, CA; Department of Management Science, and Engineering, Stanford University, Stanford, CA; Stanford University School of Medicine; Stanford, CA; Stanford Center for Biomedical Informatics Research (BMIR), Dept of Medicine and Dept of Biomedical Data Science

**Keywords:** glioblastoma, treatment patterns, unstructured data, natural language processing, abstraction

## Abstract

**Background:** Data on lines of therapy (LOTs) for cancer treatment is important for clinical oncology research, but LOTs are not explicitly recorded in EHRs. We present an efficient approach for clinical data abstraction and a flexible algorithm to derive LOTs from EHR-based medication data on patients with glioblastoma (GBM).

**Methods:** Non-clinicians were trained to abstract the diagnosis of GBM from EHRs, and their accuracy was compared to abstraction performed by clinicians. The resulting data was used to build a cohort of patients with confirmed GBM diagnosis. An algorithm was developed to derive LOTs using structured medication data, accounting for the addition and discontinuation of therapies and drug class. Descriptive statistics were calculated and time-to-next-treatment analysis was performed using the Kaplan-Meier method.

**Results:** Treating clinicians as the gold standard, non-clinicians abstracted GBM diagnosis with sensitivity 0.98, specificity 1.00, PPV 1.00, and NPV 0.90, suggesting that non-clinician abstraction of GBM diagnosis was comparable to clinician abstraction. Out of 693 patients with a confirmed diagnosis of GBM, 246 patients contained structured information about the types of medications received. Of those, 165 (67.1%) received a first-line therapy (1L) of temozolomide, and the median time-to-next-treatment from the start of 1L was 179 days.

**Conclusions:** We also developed a flexible, interpretable, and easy-to-implement algorithm to derive LOTs given EHR data on medication orders and administrations that can be used to create high-quality datasets for outcomes research. We also showed that the cost of chart abstraction can be reduced by training non-clinicians instead of clinicians.

**Importance of the study:** This study proposes an efficient and accurate method to extract unstructured data from electronic health records (EHRs) for cancer outcomes research. The study addresses the limitations of manual abstraction of unstructured clinical data and presents a reproducible, low-cost workflow for clinical data abstraction and a flexible algorithm to derive lines of therapy (LOTs) from EHR-based structured medication data. The LOT data was used to conduct a descriptive treatment pattern analysis and a time-to-next-treatment analysis to demonstrate how EHR-derived unstructured data can be transformed to answer diverse clinical research questions. The study also investigates the feasibility of training non-clinicians to perform abstraction of GBM data, demonstrating that with detailed explanations of clinical documentation, best practices for chart review, and quantitative evaluation of abstraction performance, similar data quality to abstraction performed by clinicians can be achieved. The findings of this study have important implications for improving cancer outcomes research and facilitating the analysis of EHR-derived treatment data.

## Introduction

One of the biggest impediments to cancer outcomes research is extracting data from unstructured clinical documents^1^. Unstructured data refers to free-form text, prose, or data that does not follow a consistent format when entered into the electronic health record (EHR). In cancer research, several critical variables are often found exclusively as unstructured data, such as diagnosis confirmation, date of diagnosis, biomarker test date, biomarker status, oral chemotherapies, clinical trial drugs, tumor recurrence and progression, tumor response, date of death, and more^2–5^.

Manual abstraction or chart review is a common method to extract unstructured data whereby humans — often clinicians — read through EHRs and record values of interest. Not only is this process slow and costly, but best practices for manual abstraction are often not implemented, leading to poor data quality and wasted effort^6^. While technological solutions to partially or completely automate chart review have been explored^7–9^, they still face substantial limitations in accuracy, generalizability, and interpretability, suggesting that manual abstraction will remain an important part of the research process for the foreseeable future^4,9–12^. Therefore, more efficient processes for chart abstraction are needed.

EHR-derived medication data plays an important role in clinical research. In clinical oncology, tracking the chronology of medication administrations and corresponding patient outcomes is critical for making treatment decisions. The chronology of a patient’s treatment can be captured in a “line of therapy” (LOT) – a specific treatment regimen administered to patients between defined start and end dates^13^. In practice, the precise start and end dates of LOTs may be blurred; clinicians may make changes to patients’ treatment plans based on translational medical advances, side effects of therapy, and novel therapeutic strategies. In research, however, defining precise start and end dates for LOTs is critical for conducting rigorous analyses of treatment patterns. Defining LOTs using EHR data for clinical research can be difficult because 1) start and end dates for LOTs are not explicitly documented in EHRs, 2) EHR-derived medication data typically requires substantial pre-processing to organize into LOTs, and 3) it is up to the discretion of clinical researchers on how to deal with clinical edge cases such as the addition or discontinuation of drugs to a regimen and treatment interruptions.

To address the limitations of manual abstraction of unstructured clinical data and to facilitate the analysis of EHR-derived treatment data, we developed a flexible algorithm to organize medication data for patients with glioblastoma multiforme (GBM) into LOTs and use the resulting LOT data to characterize GBM treatment patterns. GBM is the most common central nervous system malignancy, with an incidence of 3.23 cases per 100,000 people^14^. GBM patients have a median survival of only 8 months, and despite standard-of-care treatment comprising surgical resection, radiation, and chemotherapy, the cancer inevitably recurs^14–16^. Consequently, current treatments are targeted towards slowing GBM progression, reducing symptoms, and improving patients’ quality of life. With this focus on GBM disease management, there is a marked heterogeneity in treatment patterns for GBM, particularly in recurrent disease^17^. Therefore, it is imperative to develop methods for efficiently extracting LOT data on patients with GBM from EHRs.

The data used for our analysis were collected using a rigorous abstraction workflow where we trained non-clinicians (undergraduate and medical students) to perform abstraction of GBM data. We investigated whether a training program comprising detailed explanations of clinical documentation, best practices for chart review, and quantitative evaluation of abstraction performance could yield similar data quality to abstraction performed by clinicians. Taken together, we present a reproducible, low-cost workflow for clinical data abstraction and a flexible algorithm to derive LOTs from EHR-based medication data. We use the resulting dataset to conduct a descriptive treatment pattern analysis and a time-to-next-treatment analysis to demonstrate how EHR-derived unstructured data can be transformed to answer diverse clinical research questions.

## Methods

### Data source

This study was a retrospective analysis of EHR data collected at Stanford Hospital, Stanford, CA. This dataset included any patient treated at Stanford from 1998-2022. Ethics approval was granted through Stanford University IRB (#50031).

### Study design and patient population

Patients with a confirmed diagnosis of glioblastoma were identified using a combination of manual chart abstraction and machine learning. Detailed methodology is described in prior work^18^. Briefly, a cohort of 1195 patients with suspected GBM were identified by filtering pathology reports for the word “glioblastoma.” Patients younger than 18 at the date of the pathology report that contained the word “glioblastoma” were excluded. From these, confirmed GBM cases were determined through manual abstraction. This set of confirmed cases were used as training data for a machine learning model to predict confirmation of GBM diagnosis in patients’ pathology reports. The resulting machine learning model had high performance^18^: sensitivity 0.96, specificity 0.96, PPV 0.98, NPV 0.9. The modeling workflow used selective prediction, which allowed the model to identify patients whose GBM status was ambiguous — these patients were subsequently manually abstracted. In total, 693 patients with confirmed GBM diagnosis were identified. Of these, 168 patients with documented structured treatment data (medication orders and administrations) were included in the final analytic dataset. The abstraction workflow used to develop the machine learning model is described below.

### Abstraction workflow

#### Preliminary chart review

Of the 1195 patients that met the initial inclusion criteria, 637 patients (including 102 patients whose GBM status was deemed ambiguous by the machine learning model) were selected for manual abstraction to collect training data for the NLP models (Figure 1). The variables to abstract were primary diagnosis of GBM and GBM primary diagnosis date. After defining the variables and inclusion criteria, the medical student abstraction team leads (ABS & JYW) reviewed a random sample of eligible charts in parallel with the clinical lead (RT). By initially reviewing the charts internally, we developed a sense of what navigation, chart characteristics, and common language were prevalent to inform subsequent development of abstraction instructions. We then consulted our clinician lead to walk through a variety of cases, both standard and atypical, to discuss the key features that need to be addressed and captured to correctly abstract the variable of interest.

**Figure 1:**
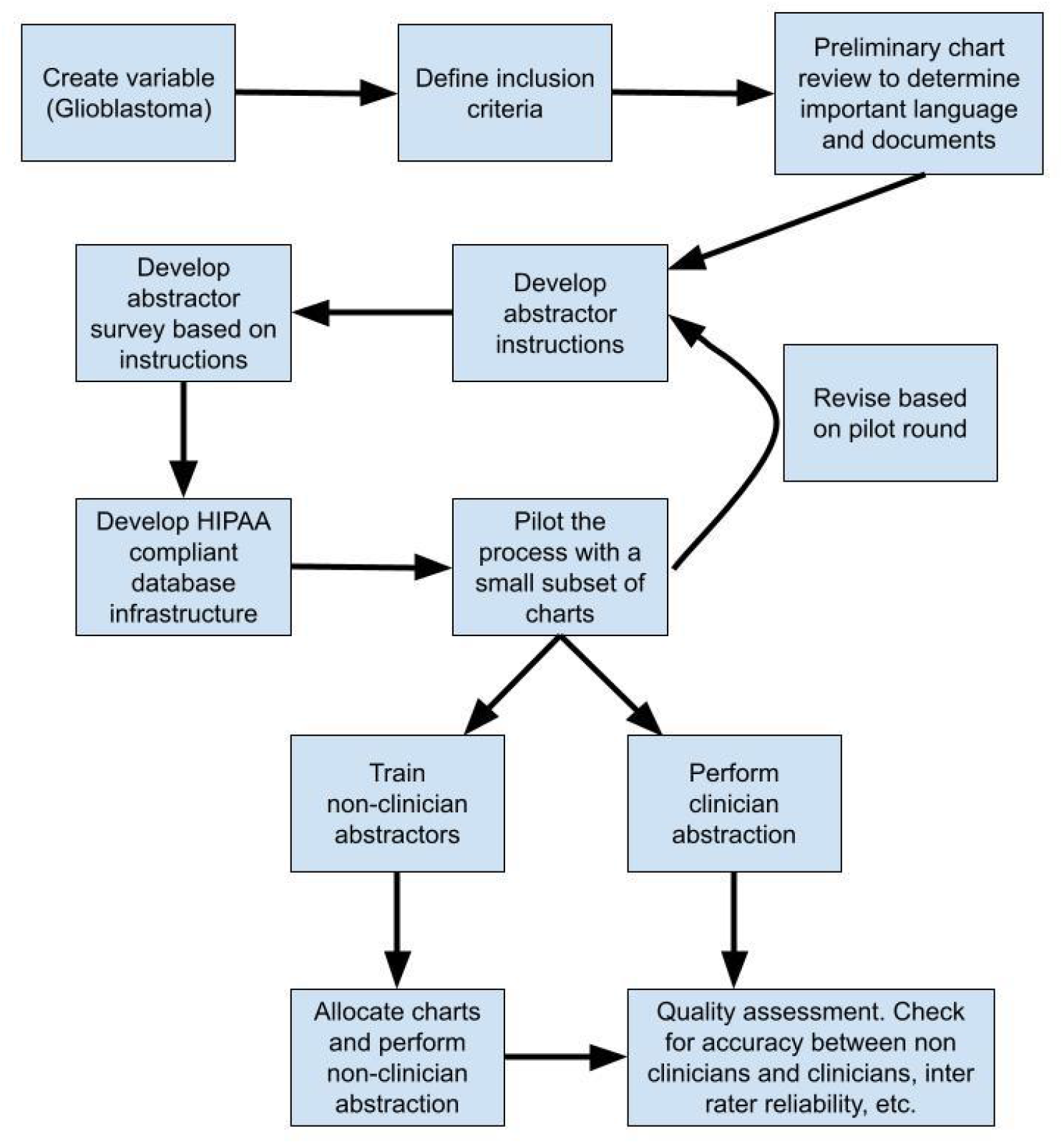
Schematic of abstraction process, starting from defining cohort inclusion criteria to performing quality assessment of abstracted data.

#### Developing abstraction instructions

After determining key considerations for clear and accurate abstractions, we created a comprehensive set of instructions designed for someone with no clinical expertise. First, we wrote a document with background information to help the non-clinical abstractor understand the disease and concepts relevant to the variable. We then defined an objective and described how to access the data and data entry tool (REDCap). Using screenshots of the data access tool and REDCap survey, the instructions explained how to navigate to the correct chart database and filter for the relevant types of pathology reports for that variable. Depending on the nature of the variable, multiple pathology reports of varying types may be present for each patient, including from procedures done at Stanford (“Surgical Procedure”) and those done externally but reviewed at Stanford (“Outside Review”). Cases with different numbers or types of charts were considered separately. The section of the chart most likely to contain the diagnosis was delineated.

#### Creating a standardized abstraction data collection tool

A REDCap survey was developed to systematically collect abstracted data using yes/no questions and drop-down menus as much as possible (SI Figure 2). This approach was utilized to maximize structured data collection, whether for the variables currently of interest or future variable abstraction. The survey included a “flag chart” function that, when selected, let abstractors choose which field in the survey was difficult to confirm. The only free-form response in the entire survey was also in this section, where we required them to elaborate on why the specific field in the survey was uncertain.

#### Pilot and revision

Student leads piloted this abstraction process, where each team member abstracted 50 charts with a 20% duplication rate for a total of 200 unique charts. The allocation and abstraction workflow otherwise followed what is described below.

This pilot round was instrumental in our abstraction development process. It illuminated inadequacies in our abstraction instructions, survey, and overall workflow. More specifically, we identified errors in our initial approach to abstracting dates of diagnosis for “outside slide review” pathology reports. Through the pilot abstraction round, we realized that the dates in the system corresponded to the dates of upload for pathology reports. These dates corresponded to surgical resection or biopsy dates on surgical pathology reports, which are accurate diagnosis dates, but for outside slide reviews, the date of specimen collection, sometimes found within the unstructured report, was instead the true diagnosis date. Discovering this error in our process to identify diagnosis dates allowed us to significantly revise our abstraction process, instructions, and REDCap survey. This step was therefore instrumental to the accuracy and quality of our abstraction process, which contributed to our creation of a more accurate dataset.

#### Abstractor training

Undergraduate abstractors were provided with basic background information on the most common types of primary central nervous tumors, grading, definitions of primary versus secondary glioblastoma, histological features of glioblastoma, and recurrence and standard of care treatment of the disease. All abstractors were required to read and familiarize themselves with this material and the extraction instructions before proceeding to the next stage of training. Subsequently, each undergraduate abstractor was assigned eight common charts (collectively referred to hereafter as a “problem set”) which they abstracted using the standardized REDCap abstraction form. These charts were specifically chosen to expose abstractors to examples of some of the most frequently encountered cases of charts that were positive or negative for confirmation of GBM diagnosis. Additionally, these charts had already been abstracted by the medical student abstraction team leads and determined to have been done so accurately by the clinician lead. Once the abstractors had completed the problem set, the medical student abstraction team leads (ABS & JYW) “graded” the submissions and then met individually with each abstractor one-on-one to walk through the solutions in detail and clarify any questions or points of confusion. ABS and JYW also created a detailed “answer key” to the problem set consisting of annotated visual diagrams of relevant chart sections, which they used both during one-on-one training sessions and subsequently provided to abstractors to use as a reference moving forward.

#### Chart allocation and abstraction

Each undergraduate abstracted 48 charts in round one with a 50% duplication rate for quality assessment. Thus, with seven undergraduate abstractors, we had 224 unique charts abstracted, with 112 duplicated. Abstractors were given one week to complete abstraction.

Afterward, the two medical student leads resolved flagged charts (in consultation with the clinician lead when necessary) to complete this first round of abstraction fully. During abstraction, non-clinicians had the option to “flag” charts where the result was uncertain. Flagged charts were reconciled by collaborative review, and uncertain cases were reviewed with a clinician.

This second round of abstraction consisted of charts that the model could not accurately predict. These remaining charts were evenly distributed among undergraduate abstractors. Abstractors were again given one week to complete their assignment, and medical student leads resolved any flagged charts.

#### Development and validation of line of therapy rules

An algorithm was developed to transform structured medication orders and administration data (one row per administration or order) into LOTs (one row per LOT). Raw structured medication data contained a unique patient identifier, the name of the drug, the date of drug administration, the date of drug order, and the expected start date of the drug. The desired data model for the resulting LOT table contained a unique patient identifier, the LOT number, the treatments included in the LOT, and the start and end dates of the LOT (SI Figure 1).

We adapted previously proposed rules for deriving LOTs from EHR data^19^. All antineoplastic drugs with a documented administration or order after the date of GBM diagnosis were included in the LOT table. Here, we define antineoplastic as including all chemotherapy, biologic, targeted therapy, or immunotherapy agents used in cancer treatment. We defined the drug use date as the first non-missing value of either the administration date, expected start date, or order date (Figure 2).

**Figure 2:**
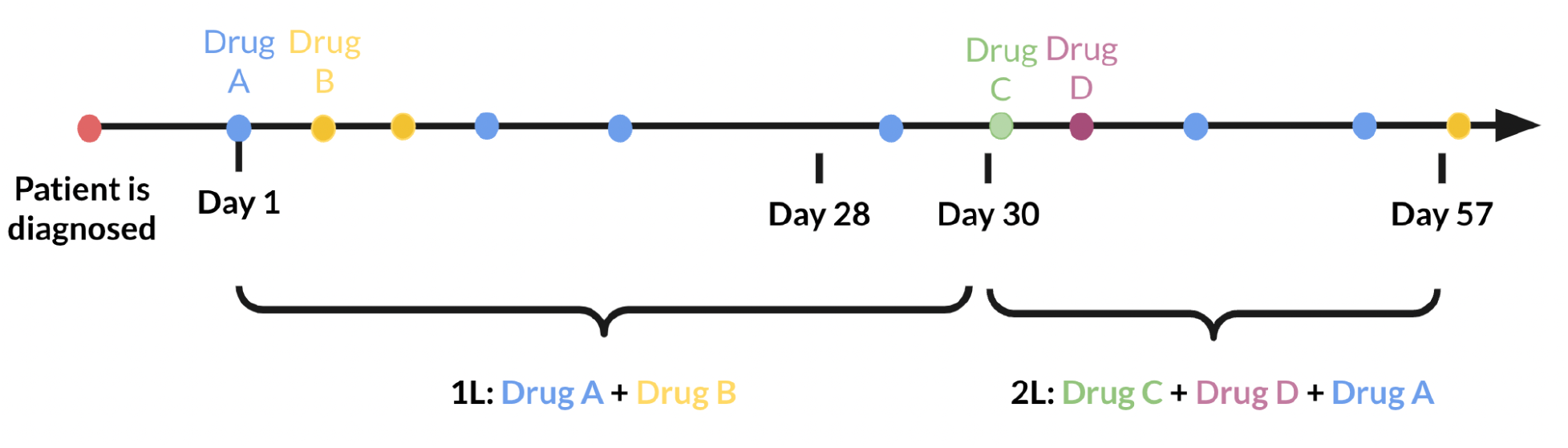
Schematic showing the definition of lines of therapy (LOT) from uses of antineoplastic drugs for an example patient. The start date of each LOT was determined based on the use date of the antineoplastic drugs, with the end date of a LOT defined as the day before the start date of the subsequent LOT. The start of the first LOT (1L) was defined as the first antineoplastic drug use date. The second LOT (2L) start date was defined as the first drug use date of a new antineoplastic not administered during the first 28 days of 1L. The 1L end date was defined as the day before 2L start.

The start of the first LOT (1L) was defined as the first antineoplastic drug use date. The second LOT (2L) start date was defined as the first drug use date of a new antineoplastic not administered during the first 28 days of 1L. The 1L end date was defined as the day before 2L start. The same logic was applied to subsequent LOTs. LOTs that are not followed by another LOT were not assigned an end date. We also defined a treatment gap rule — a rule that triggers the start of a new line when at least 90 days pass between drug uses. After applying these rules to generate a LOT table, the resulting output was reviewed by a neuro-oncologist (RT) to ensure alignment with expected treatment patterns.

## Statistical analysis

### Analysis of abstractor performance

Inter-rater reliability for abstraction of GBM diagnosis (y/n) was calculated among 112 charts duplicate-abstracted by non-clinicians using Fleiss’ kappa. Fleiss’ kappa is a measure of inter-rater reliability for a fixed number of raters classifying items, with a kappa of 1 indicating complete agreement, and a kappa ≤ 0 indicating no agreement among raters beyond what would be expected beyond chance. A kappa of ≥ 0.8 was considered superior reliability. In addition, the performance of non-clinician abstractors was compared to that of clinician abstractors using sensitivity, specificity, negative predictive value, and positive predictive value, treating clinician abstractors as the gold standard.

### Treatment patterns analysis

Descriptive statistics including most common LOTs were calculated. In the overall population, median time to next treatment (TTNT) was estimated using the Kaplan Meier estimator. The index date was the date of treatment initiation (1L), and the event date was defined as the start date of the subsequent LOT (2L). Patients who only had one LOT were censored at their last drug use date or their date of death, whichever was later.

## Results

### Cohort abstraction

A total of 659 charts were manually abstracted by non-clinicians, of which 118 were duplicate-abstracted by non-clinicians and 54 were duplicate-abstracted by clinicians. Inter-rater reliability analysis yielded a Fleiss’ Kappa of 0.869 (p-value < 0.001), indicating excellent reliability (CITE Fleiss Kappa interpretation) among non-clinicians. Treating clinicians as the gold standard, non-clinicians abstracted GBM diagnosis with sensitivity 0.98, specificity 1.00, PPV 1.00, and NPV 0.90, suggesting that non-clinician abstraction of GBM diagnosis was comparable to clinician abstraction.

### Cohort Description of Lines-of-Therapy

Following our model-assisted abstraction process, we assembled a cohort of 693 patients with a confirmed diagnosis of glioblastoma. From that group, 246 patient charts contained structured information about the types of medications received. Using medication data, we generated lines of therapy using a custom set of rules, specifically filtering for antineoplastic drugs (Table 1). Of the 246 patients, 165 (67.1%) received a first-line therapy (1L) of temozolomide, 33 patients (13.4%) received bevacizumab alone and 14 patients (5.7%) received a combination of bevacizumab and temozolomide.

**Table 1:**
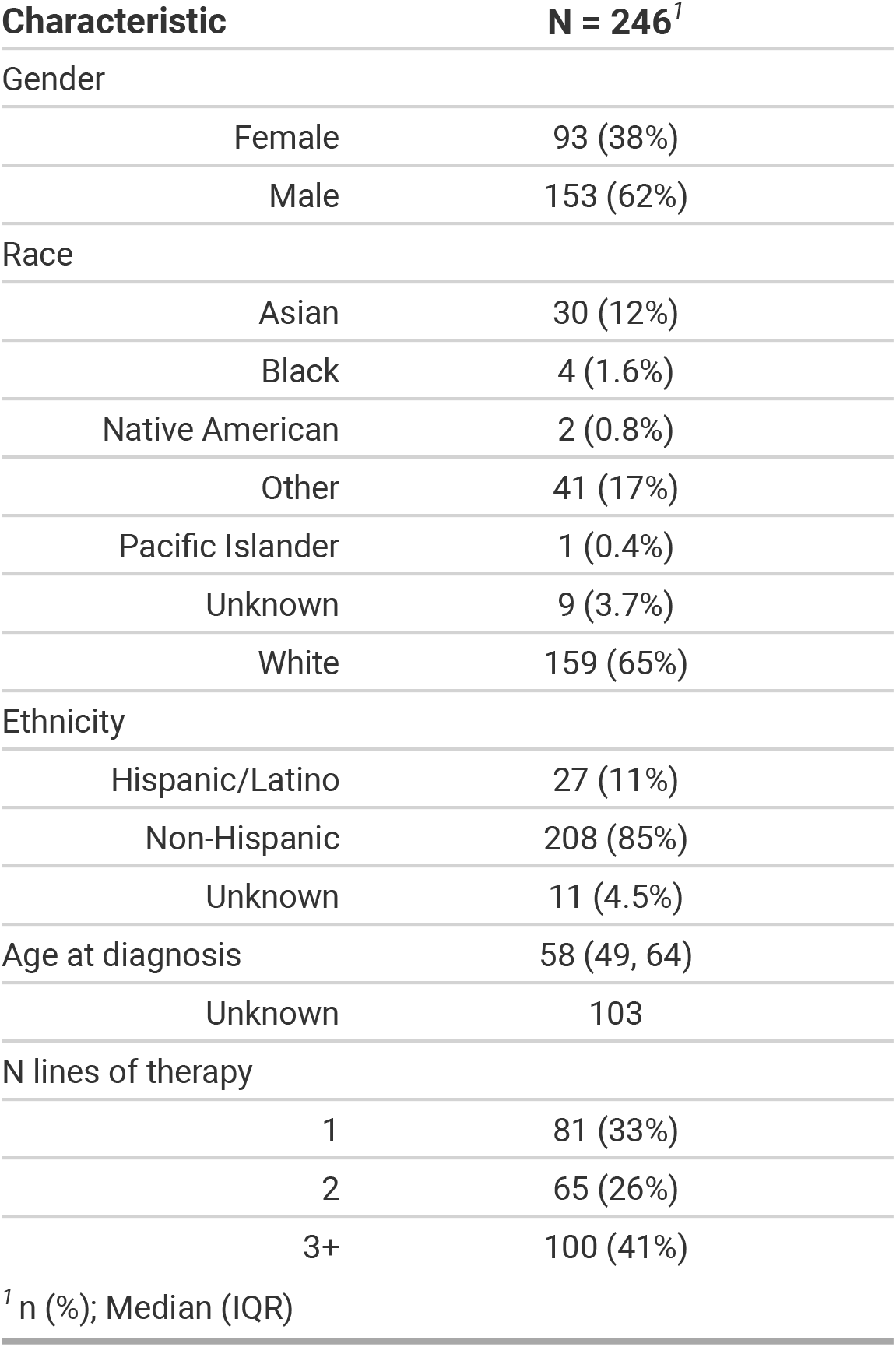
Demographic and clinical characteristics of the 246 patients receiving treatment for GBM.

Among the 246 patients with a documented 1L, 165 patients (67.1%) progressed on to a second-line therapy (2L). Progression from 1L to 2L was analyzed for each type of treatment in a Sankey diagram (Figure 3). 165 patients received temozolomide in 1L compared to 40 patients in 2L. 33 patients received bevacizumab in 1L compared to 55 patients in 2L. 14 patients received a combination of temozolomide and bevacizumab in 1L compared to 22 patients in 2L.

**Figure 3:**
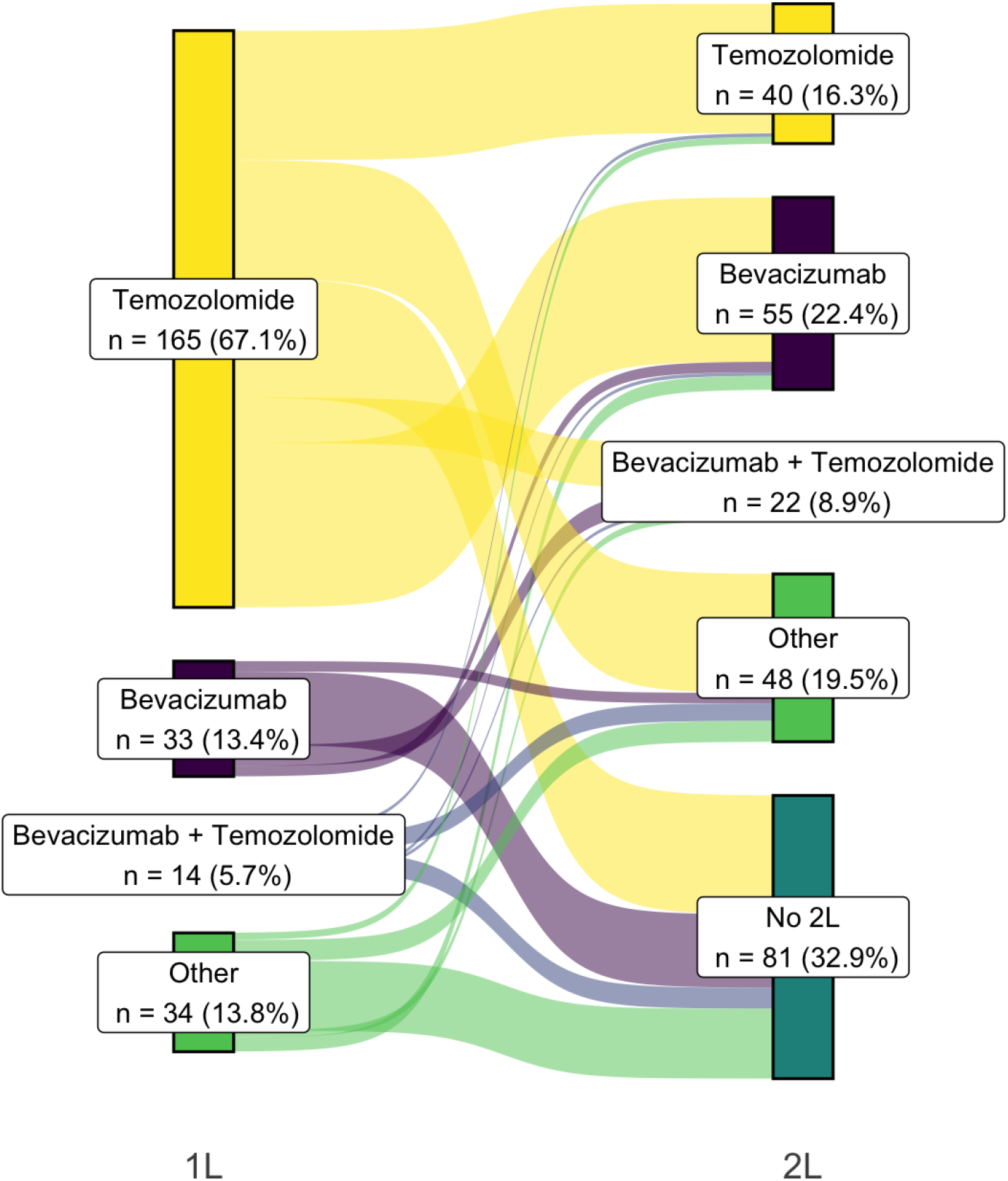
Sankey diagram showing the progression of 1L to 2L treatments.

**Figure 4.**
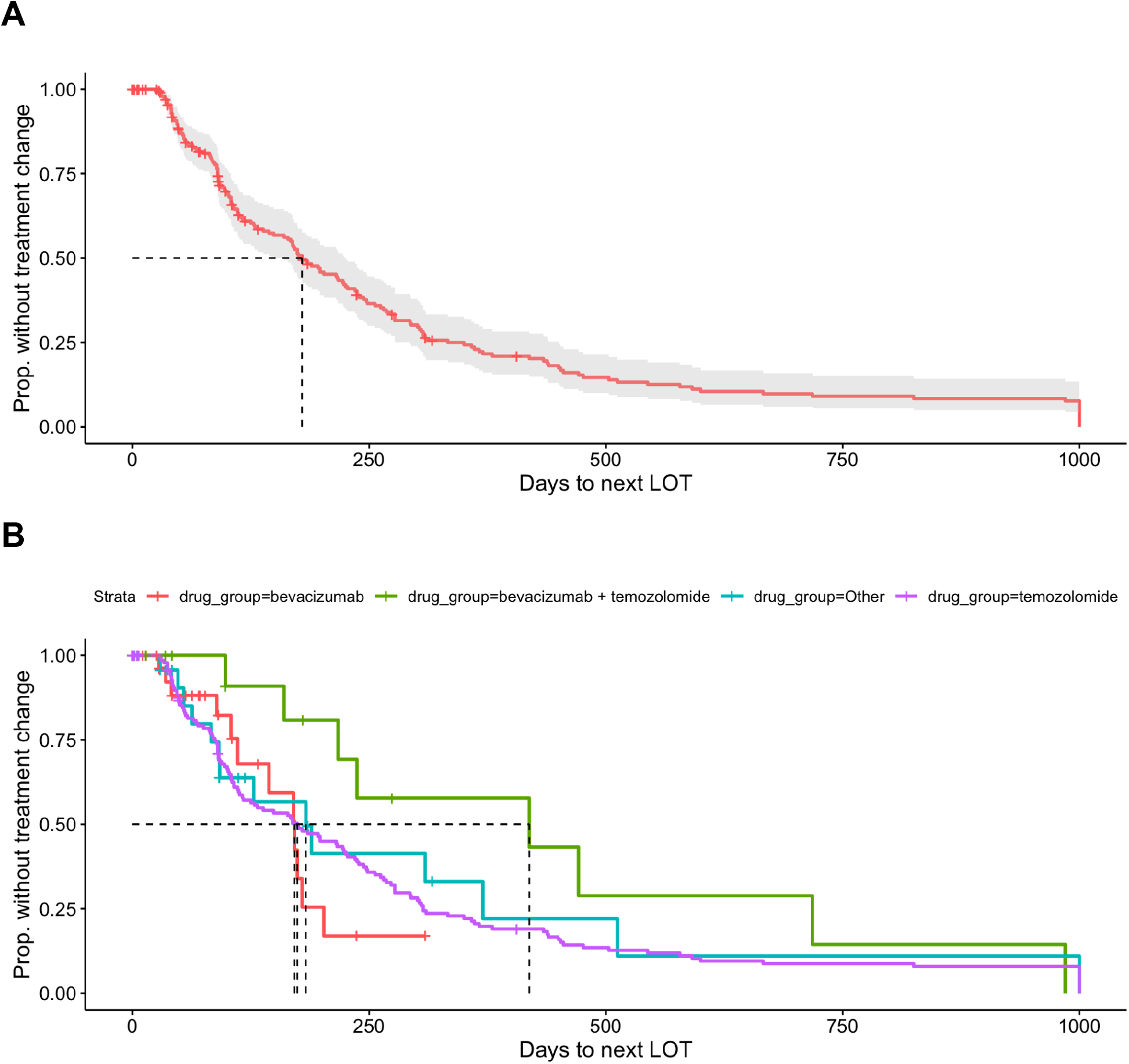
Kaplan Meier curves plotting time-to-next-treatment (TTNT) for the entire cohort of 246 GBM patients and their 1L treatment regimens (**Panel A**), as well as comparisons between 1L Temozolomide, 1L Bevacizumab, 1L Bevacizumab + Temozolomide and all other treatments (**Panel B**). Y-axis represents proportion of patients continuing current treatment and x-axis represents days since treatment initiation. Grey and multicolored dotted regions represent 95% confidence intervals (where available).

### Time-to-Next-Treatment Analysis for First-Line and Second-Line Therapies

The median time-to-next-treatment (TTNT) for all 1L treatments was 179 days (95% CI 149-225) (Table 2). Among the 165 patients who received 1L temozolomide, the median TTNT was 174 days (95% CI 117-227). Among the 33 patients who received 1L bevacizumab, median TTNT was 171 days (95% CI 111-NA). Among the 14 patients who recieved 1L temozolomide and bevacizumab combined, median TTNT was 419 days (95% CI 217-NA). The median TTNT across all 2L treatments was 195 days (95% CI 146-242). Among patients who received 2L temozolomide, median TTNT was 206 days (95% CI 113-445). Among patients who received 2L bevacizumab, median TTNT was 195 days (95% CI 122-823). Among patients who received 2L temozolomide and bevacizumab combined, median TTNT was 225 days (95% CI 163-874).

**Table 2.** Descriptive statistics of TTNT analysis for primary 1L and 2L treatments. Median TTNT (duration) for bevacizumab and temozolomide combination is much greater than other regimens for first and second LOTs. Nonetheless, temozolomide is the most common 1L treatment while bevacizumab is the most common 2L.

| Line | Treatment | N | % | Median duration (days) | 95% LCL | 95% UCL |
| --- | --- | --- | --- | --- | --- | --- |
| 1L | All Treatments | 246 | 100% | 179 | 149 | 225 |
| 1L | temozolomide | 165 | 67.1% | 174 | 117 | 227 |
| 1L | Other | 34 | 13.8% | 183 | 92 | NA |
| 1L | bevacizumab | 33 | 13.4% | 171 | 111 | NA |
| 1L | bevacizumab + temozolomide | 14 | 5.7% | 419 | 217 | NA |
| 2L | All Treatments | 165 | 100% | 195 | 146 | 242 |
| 2L | bevacizumab | 55 | 33.3% | 195 | 122 | 823 |
| 2L | Other | 48 | 29.1% | 119 | 80 | 267 |
| 2L | temozolomide | 40 | 24.2% | 206 | 113 | 445 |
| 2L | bevacizumab + temozolomide | 22 | 13.3% | 225 | 163 | 874 |

## Discussion

Unstructured EHR data contains valuable information for outcomes research, but is difficult to analyze in its raw form. Often, highly trained human abstractors (e.g. clinicians) are required to manually extract unstructured data from EHR records, which is expensive and time-consuming. Here, we presented two methods of extracting and processing unstructured clinical data. First, we described a rigorous approach to train non-clinician abstractors to extract clinical data on patients with GBM, and showed that the accuracy of non-clinician abstractors was comparable to that of clinician abstractors. This approach could be used to reduce the cost of abstraction by leveraging non-clinicians instead of clinicians. Second, we developed a flexible, interpretable, and easy to implement algorithm to derive LOTs given EHR data on medication orders and administrations.

Our analysis of the derived LOTs showed that our cohort’s treatment patterns aligned with the typical standard of care for GBM. We compared our cohort to other GBM cohorts with well-described treatment patterns and found that our cohort had similar characteristics in terms of LOT agents and durations. For instance, Girvan et al. assembled a cohort of 503 GBM patients using an online chart abstraction process conducted by 160 participating oncologists. Their cohort predominantly received 1L temozolomide monotherapy (76.5%) and 2L bevacizumab monotherapy (58.1%), and the median 2L duration was 130 days^20^. In comparison, the most common 1L and 2L in our cohort were temozolomide monotherapy (67.1% of patients receiving 1L therapy) and bevacizumab monotherapy (33.3% of patients receiving 2L therapy), and the median 2L duration was 195 days. However, we note that all patients included in the Girvan cohort received at least two LOTs, whereas only 67.1% of patients in our cohort received 2L therapy. Another cohort of 750 GBM patients assembled by Annavarapu et al. showed that the majority of patients received 1L radiation concurrent with temozolomide (90.1%), bevacizumab was the most common 2L therapy (73.4% of patients receiving 2L therapy), and the median 1L duration was 135.1 days^21^. Annavarapu et al. defined treatment duration as the interval between the date of LOT initiation and the date of last administration, which therefore does not account for the date of initiation for the subsequent LOT. Given that our GBM cohort’s treatment features were broadly similar to those of other published cohorts, we believe that our abstraction process and LOT algorithm reasonably reflected GBM patients’ treatment journeys.

Our approach has some limitations. First, since our GBM cohort was assembled in part using a machine learning model, there could have been false positives (patients who did not truly have GBM) in the cohort. However, this would have been a rare occurrence given the strong performance of the machine learning model on a naive test set. Second, the sample size used for the LOT analysis was small due to the limited number of patients with documented treatment in the EHR. Third, our LOT algorithm uses a simple set of rules that may not capture all clinical nuances in lines of therapy. Further work may build upon this initial algorithm by conducting more in depth validation and expanding the rule set to account for more clinical nuances.

Overall, our workflow for unstructured data extraction using non-clinician abstractors and the customizable LOT algorithm are important contributions that can be applied to a wide range of clinical research scenarios. Utilizing LOTs from a large dataset of GBM treatments as a tool for determining outcomes of these different treatment options can pave the way for identifying the most effective regimens that maximize patient survival. LOT analysis can provide insights into the overall survival, progression-free survival, and time to next treatment of specific regimens and their corresponding modalities. Such information will be crucial towards formulating new standards of care that utilize every available resource to improve patient outcomes.

## Data Availability

All data needed to evaluate the conclusions are present in the paper and in the Supplementary Materials.
The datasets generated analyzed during the current study are not publicly available due to patient privacy but are available from the corresponding author (AS) on reasonable request.

## Supplementary Information

**SI Fig. 1.**
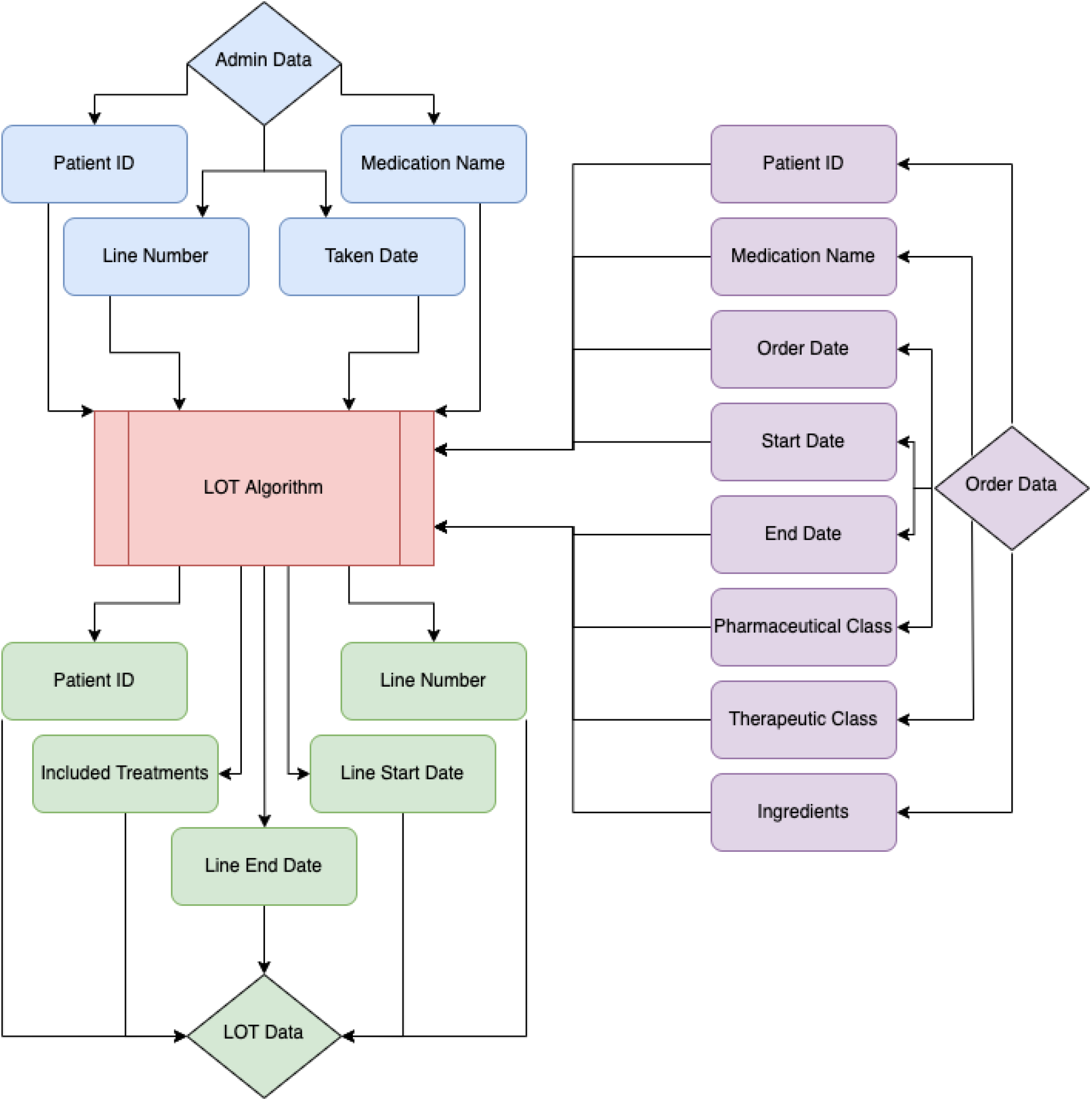
Schematic diagram of line of therapy (LOT) algorithm inputs and outputs.

**SI Figure 2:**
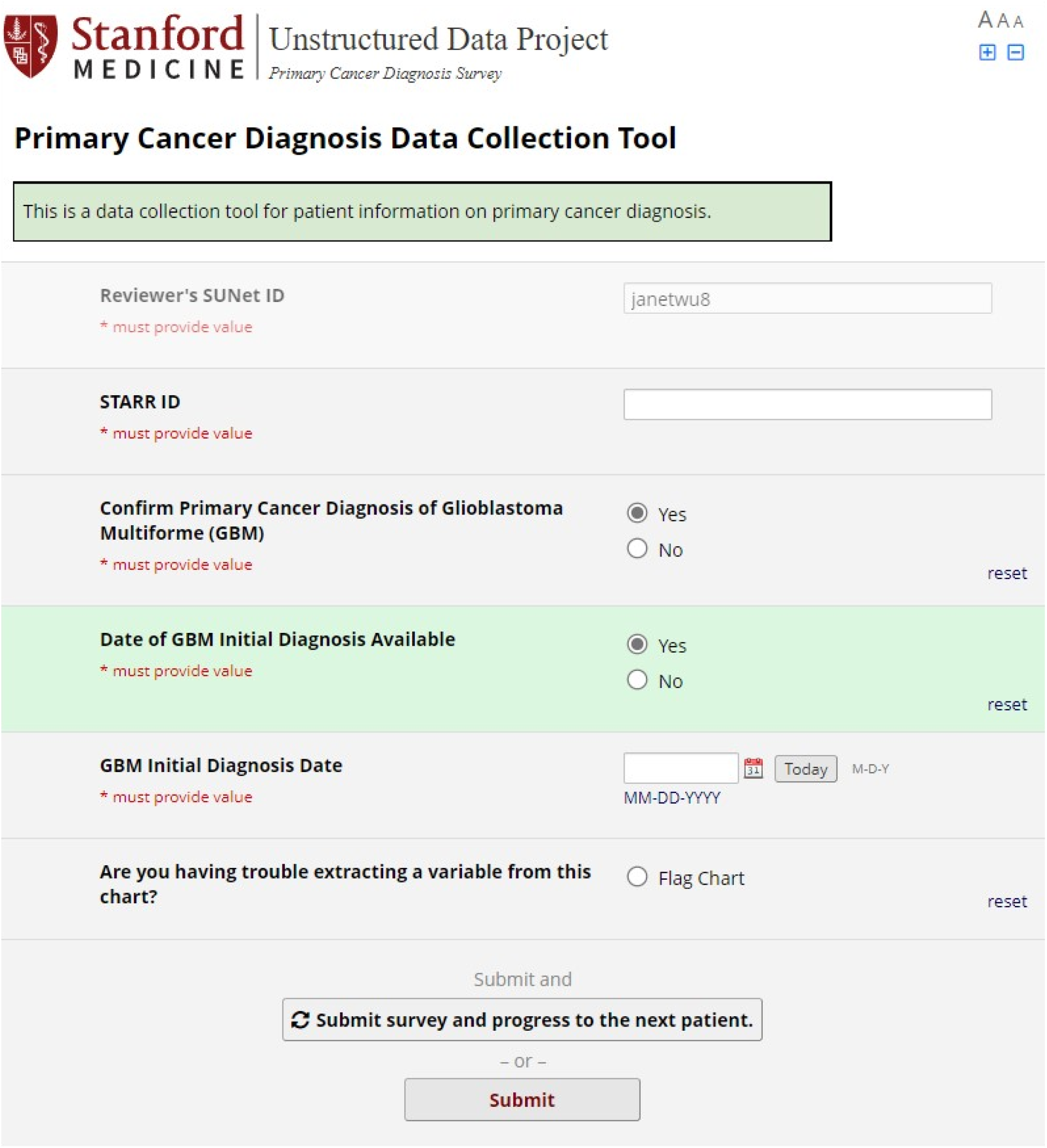
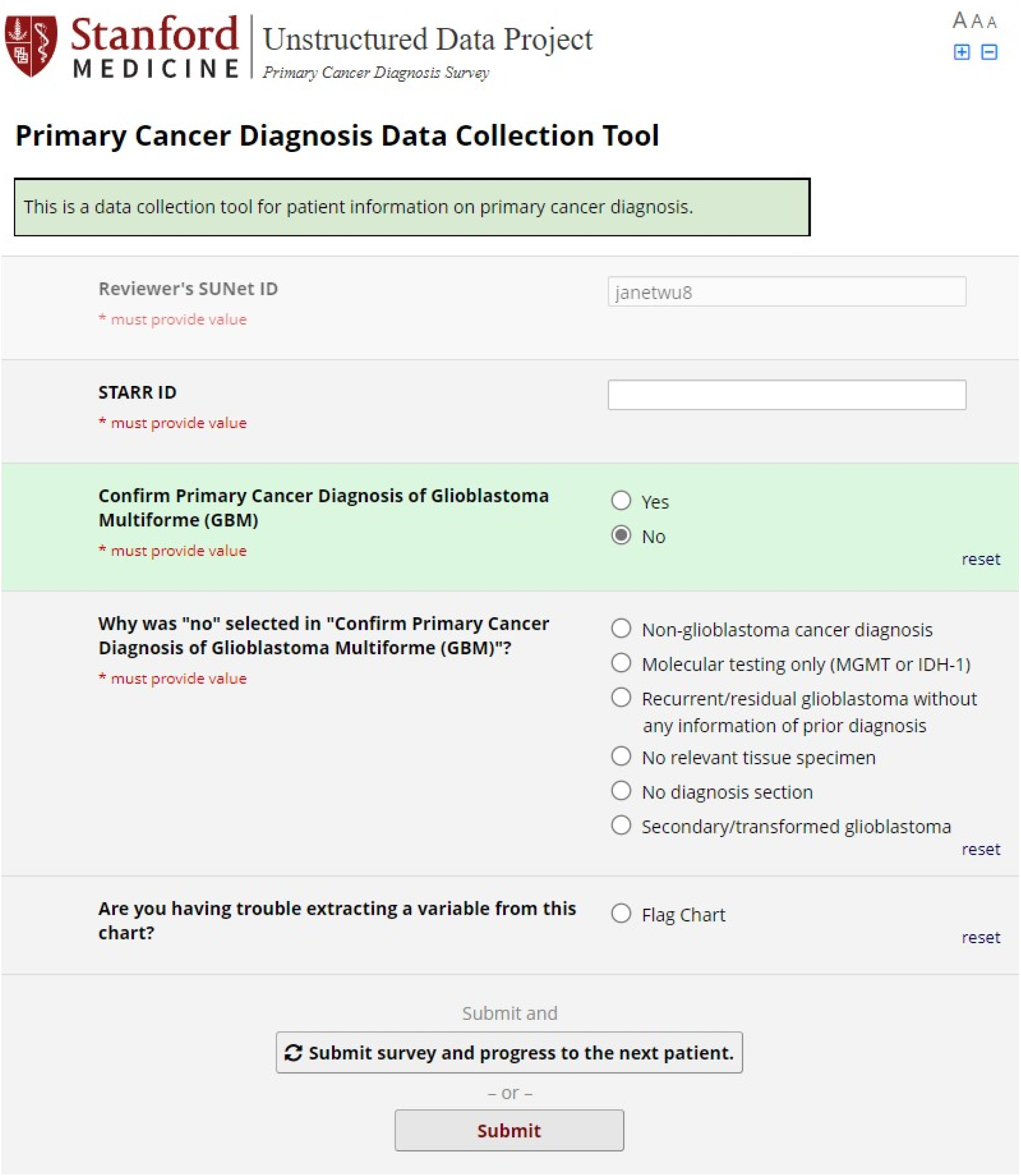
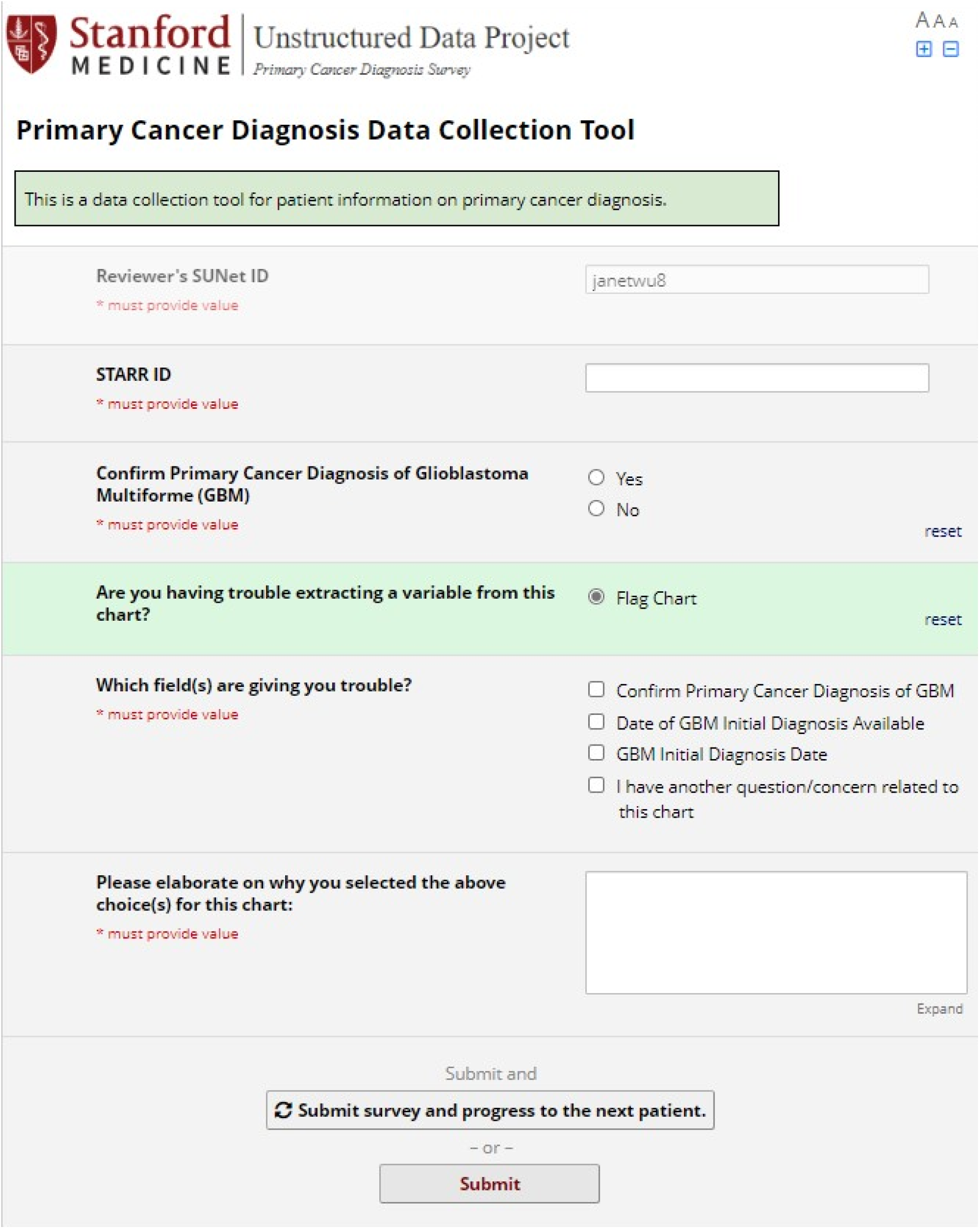
Screenshots of the abstraction data collection tool

